# A scalable and equitable framework for target and patient prioritisation in rare disease antisense therapeutics

**DOI:** 10.64898/2026.03.23.26348690

**Authors:** Ella F. Whittle, Kylie-Ann Montgomery, Carme Camps, Nour Elkhateeb, Catherine Ryan, Sara Aguti, Thales A. C. de Guimarães, Usha Kini, Helen Stewart, Andrew G. L. Douglas, Louise Wilson, Harry G. Leitch, David S. Lynch, Robert Robinson, Michel Michaelides, Timothy W. Yu, Paul Gissen, Marlen C. Lauffer, Nick Lench, Dan O’Connor, Ana Lisa Tavares, Stephan J. Sanders, Manju A. Kurian, Hannah Titheradge, Emma Clement, Jacqueline van der Spuy, Jenny C. Taylor, Carlo Rinaldi, Francesco Muntoni, Haiyan Zhou, Alice E. Davidson, Mina Ryten, UPNAT consortium

**Author notes:** These authors contributed equally (Kylie-Ann Montgomery, Carme Camps, Nour Elkhateeb, Catherine Ryan). **Corresponding authors with equal contributions**.

## Abstract

**Background:** Nucleic acid therapies (NATs) comprise engineered DNA- or RNA-based medicines that act through sequence-specific interactions to modify gene function. Among these, antisense oligonucleotide (ASO) therapies are designed to bind messenger RNA (mRNA) or pre-mRNA to alter splicing, transcript stability, or translation. Many patients with a rare genetic disease stand to benefit from these treatments and, as underlying technologies continue to advance, a critical barrier to care is the equitable selection of targets and patients. Owing to landmark progress in genomic health care, the UK is uniquely positioned to develop a national framework on NAT patient-selection infrastructure. The UK Platform for Nucleic Acid Therapies (UPNAT) has been launched, in part, to meet this goal, with a key output being a structured patient and target selection framework to support NAT development and clinical application, using ASO therapies as a pilot modality.

**Methodology and Results:** A multidisciplinary panel of UK-based experts established the UPNAT framework to enable systematic assessment of ASO amenability across modular domains encompassing disease understanding, functional models, variant characteristics, and the individual patient, incorporating the recently published N1C VARIANT guidelines. This modular structure supports consistent prioritisation of tractable targets while identifying biological, clinical, technical, or evidentiary gaps currently limiting ASO development. Designed for implementation within the UK healthcare infrastructure and amenable to future automation using open-access resources, the framework was iteratively refined through application to genomic and clinical data from approved ASO therapies and selected real-world patient case studies.

**Conclusion:** We present the first disease-agnostic framework to support structured prioritisation of patients and targets (diseases, genes, or variants) for ASO development and consideration within specialist healthcare services. Designed to accommodate rapid technological advances in NATs, the framework promotes transparent, equitable, and reproducible decision-making within the UK National Health Service (NHS), with principles transferable to other healthcare systems.

## Introduction

Rare genetic diseases collectively represent a substantial global challenge, affecting approximately 300 million individuals worldwide and placing sustained strain on patients, families, and healthcare systems (1). Although advances in genomic diagnostics have transformed the ability to identify rare disease aetiologies, therapeutic options remain limited, particularly for ultra-rare and genetically heterogeneous conditions. Antisense oligonucleotides (ASOs) have emerged as a promising class of precision nucleic acid therapies (NATs), offering a flexible means to modulate gene expression through diverse mechanisms, including splice modification, transcript degradation, suppression of toxic interactions, or upregulation of gene expression (2–7). This mechanistic flexibility enables ASO strategies that are gene-or variant-specific, variant-agnostic (for example, targeting common cis-acting variants), or directed towards compensatory pathways, thereby expanding the range of patients who may be eligible for therapeutic development. As ASO technologies mature and their clinical feasibility is increasingly demonstrated, this expansion in therapeutic possibility creates a new translational bottleneck: determining which patients and targets (diseases, genes, or variants) should be prioritised for ASO development and treatment in a manner that is equitable, transparent, and scalable within healthcare systems.

This decision pressure is no longer theoretical. Clinical translation of ASO therapies has accelerated in recent years, encompassing both commercially approved drugs and compassionate-use treatments developed for individual patients (**Figure 1A**). Available data demonstrate a clear divergence in development strategies: ASOs designed for a specific individual predominantly adopt variant-specific approaches, whereas therapies achieving full market approval more commonly employ non-variant-based strategies that can be applied across broader patient groups (**Figure 1B**). This capacity to tailor treatment to the root genetic cause underpins the success of ASOs as N-of-1 therapeutics, exemplified by the development of Milasen in 2018 (8). However, the same flexibility that enables individualised and more broadly applicable ASO strategies also multiplies the number of plausible development pathways, candidate patients, and decision points, intensifying the challenge of prioritisation in the absence of structured guidance.

**Figure 1:**
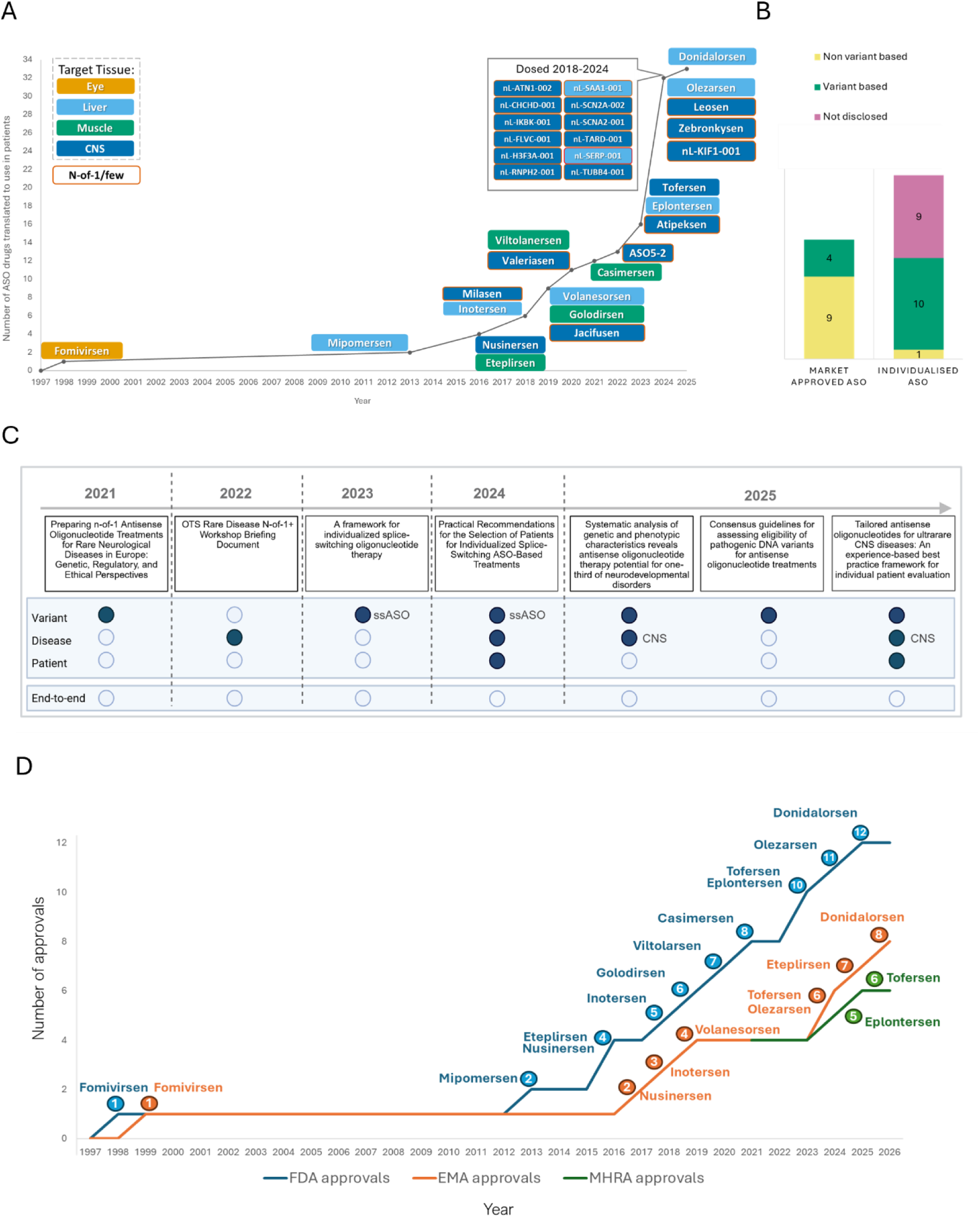
The growing urgency for end-to-end guidance for patient prioritisation in light of increased clinical translation of ASO-based therapies for rare disease. **A)** Market approved and N-of-1/few individualised ASO therapies. Dates reflect year of approval (by the FDA, EMA or MHRA) or first use in patients. If market approved, the date reflects the earliest date, regardless of the regulatory body. It should be noted that Viltolarsen was first approved by the regulatory system in Japan. **B)** Distinction between variant strategy (variant specific vs variant agnostic) across ASO therapies reported in clinical use (Figure 1A). Market approved ASO therapies on the left and individualised ASO therapies that are, or have been, in clinical use until 2024 shown on the right (13). **C**) Recommendations available to guide patient selection for ASO therapies (10, 14–19). Year of guidance release is indicated along the arrow. Three main focal areas are covered: variant, disease, and patient. Lacking is end-to-end guidance readily implementable within the UK NHS. **D)** Rate of market ASO approvals since the first ASO approval in 1998 by the FDA, EMA and post 2021 MHRA.

Against this backdrop, a critical barrier to clinical translation of ASO therapies is the equitable and consistent selection of appropriate patients and targets, an issue increasingly recognised across gene-based therapeutic modalities (9). Transparent guidance is therefore required to support systematic prioritisation and timely progression from bench to bedside. Although ASO mechanisms are versatile, not all genetic variants are amenable to ASO intervention, a limitation addressed at the variant level by the N=1 Collaborative (N1C) VARIANT consensus guidelines for assessing eligibility of pathogenic DNA variants for ASO treatments (10). However, variant eligibility alone is insufficient to guide clinical decision-making. Not all genes, diseases, or individual patients are suitable candidates for ASOs, and existing guidance does not integrate disease biology, functional model readiness, patient-level risk–benefit considerations, or health-system feasibility into a unified assessment. While a growing body of literature offers expert recommendations for ASO patient selection (**Figure 1C**), no disease-agnostic, end-to-end framework currently exists, nor is there UK-specific guidance aligned with the regulatory and structural constraints of the National Health Service (NHS).

The absence of such a framework is particularly consequential in the UK. As a world leader in genomic healthcare and a pioneer of national diagnostic and research programmes, the UK is uniquely positioned to deliver NATs within a publicly funded health system. More than 3.5 million people in the UK live with a rare disease, and national initiatives are rapidly expanding access to genomic diagnosis. Programmes and databases such as the Genomics England Generation Study and Genomics England National Research Library are improving diagnoses and shifting diagnosis to earlier stages of life, increasing the number of individuals identified at a point where therapeutic intervention may be feasible or most effective. This expanding denominator, combined with distinct regulatory and resource constraints, intensifies the need for transparent, equitable, and reproducible approaches to prioritisation. The UK Platform for Nucleic Acid Therapies (UPNAT), funded by the National Institute for Health and Care Research (NIHR) and Medical Research Council (MRC) and aligned with initiatives such as the Rare Therapies Launchpad (RTLP) and MRC Centre of Research Excellence (CoRE) in Therapeutic Genomics, was established to address this challenge. Here, we present the first disease-agnostic patient selection framework, using ASOs as a pilot modality, for initial application within the UK. Within this evolving implementation landscape, the Medicines and Healthcare products Regulatory Authority (MHRA) has held independent responsibility for pharmaceutical approvals in the UK since 2021 and has recently outlined initiatives aimed at improving pathways for innovative and rare disease therapies. Differences in the pace of ASO therapy approvals relative to other jurisdictions (**Figure 1D**) highlight the importance of complementary decision frameworks that can help translate technological opportunity into clinical implementation. With updated clinical trial legislation intended to streamline regulatory pathways and support access to innovative treatments (11, 12), the UK provides an important setting for responsible development of novel NATs.

By separating biological feasibility from clinical and system-level prioritisation, the framework supports clear and reproducible decision-making. The framework is designed to promote equity, transparency, and scalability by enabling consistent assessment across diverse diseases, therapeutic strategies, and clinical contexts. It integrates four modular dimensions - disease biology, functional model readiness, variant amenability, and patient-level considerations - within a unified assessment structure. By distinguishing biological feasibility from clinical and system-level prioritisation, the framework supports reproducible decision-making applicable to both generalisable and individualised ASO strategies. Specialist expertise is embedded throughout the process in a structured and comparable manner across disease areas. The governance model is tiered: disease and functional model assessments require episodic, centralised domain-specific evaluation with reusable outputs, whereas patient-level assessment is designed for iterative local application. This structure mirrors established UK genomic governance models, in which centralised determinations of eligibility are coupled with locally delivered clinical implementation under licensed oversight frameworks.

In this study, we describe the development of this framework through multidisciplinary expert consensus and its calibration using targets of market-approved ASO therapies. We then demonstrate its real-world applicability through assessment of patients considered for ASO development, illustrating how the framework supports both prioritisation and principled exclusion where appropriate based on current knowledge. Together, these analyses show how structured patient selection can translate expanding genomic and therapeutic capability into consistent, equitable decision-making, providing a model for the responsible deployment of precision NATs within the UK NHS, based upon core principles generalisable to other healthcare systems.

## Materials and methods

### Scope and objectives

The UPNAT Target Selection Framework is designed to support structured decision-making for ASO therapies. Specifically, it provides a framework for: (i) assessing patient suitability for treatment with existing ASO therapies; (ii) determining whether specific disease–gene targets are appropriate candidates for novel ASO development; and (iii) identifying key biological, clinical, technical, or evidentiary gaps that currently limit therapeutic development. The framework is applicable to both individualised (variant-specific) and generalised (gene- or pathway-directed) ASO strategies within rare genetic disease.

The framework focuses on biological tractability, current technical feasibility and clinical appropriateness, using explicit exclusion and prioritisation criteria to support consistent assessment across diverse diseases and clinical contexts. It is intended to inform early-stage triage and prioritisation decisions rather than late-stage clinical trial readiness. Of note, regulatory considerations surrounding therapy development and reimbursement or financial delivery models are important factors that are beyond the scope of this initial framework, which are addressed by parallel and complementary initiatives, such as the Rare Therapies Launchpad UK pilot program (20).

Our objectives were to:

1. Promote equitable patient selection by applying transparent and reproducible criteria, ensuring that all patients are assessed on a common basis within the NHS.
2. Enable consistent and timely prioritisation of disease–gene targets for ASO development based on biological, clinical, and technical feasibility.
3. Support clear and efficient decision-making by providing structured guidance that can be applied at defined points in the translational pathway.
4. Structure assessment to ensure specialist expertise is applied where most critical, with local implementation guided by predefined, specialist-developed criteria.

### Framework consensus development process

The UPNAT Target Selection Framework was developed through a structured, iterative consensus process involving multidisciplinary experts in rare disease, NATs, clinical genetics, and translational medicine (**Supplementary Figure 1**). An initial multi-stakeholder meeting was convened to define the scope and guiding principles of the framework and to identify key domains required for patient and target selection. From this meeting, a core development group was established (**Supplementary Table 1**) and organised into four module-specific Working Groups corresponding to disease, functional model, patient, and variant assessment.

Each Working Group was provided with a standardised module template outlining required decision points, exclusion criteria, and scoring principles. Module leads were responsible for drafting and refining criteria within their assigned domain, with iterative feedback exchanged within and between groups. Draft modules were reviewed collectively at an in-person workshop, where criteria definitions, scoring structure, and implementation considerations within the UK healthcare system were discussed and refined. Subsequent whole-group meetings were used to incorporate feedback arising from pilot application of the framework and to resolve areas of ambiguity.

This framework was developed as a pragmatic decision-support tool rather than a consensus guideline, with a focus on UK-based implementation; accordingly, formal Delphi-style consensus scoring was not employed, as the emphasis was on reproducibility, rather than convergence of expert opinion alone. Where differences of opinion arose, criteria were retained only when agreement could be achieved across both clinical and research assessors, ensuring that decisions reflected feasibility within real-world clinical pathways as well as biological plausibility. Transparent and iterative communications were key to framework development and thus expert consensus was developed through non-anonymised structured multidisciplinary discussion.

Framework development and refinement occurred over an approximately 12-month period, allowing iterative revision informed by pilot application and cross-disciplinary review. Criteria and scoring frameworks were finalised prior to systematic application to market-approved ASO targets and patient case examples. The resulting framework integrates clinical, research, and translational perspectives and was explicitly designed to balance biological feasibility with practical implementation within NHS clinical pathways.

### Testing on market approved ASO targets

To assess whether the disease and functional model modules of the UPNAT Target Selection Framework were appropriately specified, we applied the framework to a set of market-approved ASO therapy targets as positive controls (**Supplementary Table 2**). This analysis was designed to calibrate the framework against targets with established clinical tractability, rather than to retrospectively validate predictive performance.

For each therapy, the disease indication and gene target were independently evaluated by multiple assessors using the relevant framework modules. Eleven ASO therapies, representing eight distinct gene–disease pairs, were included. Therapies were selected if they targeted monogenic, non-infectious disorders with well-characterised Mendelian architecture and established oligonucleotide chemistry backbones with known safety profiles. While publicly available data were sufficient to support assessment of targets that progressed to clinical use, comparable information for ASO programmes that were discontinued or deemed non-targetable was not available. As a result, this analysis was not intended to establish sensitivity or specificity, but rather to confirm that framework module criteria did not inappropriately exclude targets that have reached clinical use.

### Case examples of ASO patient assessments

To demonstrate the real-world usability of the UPNAT Target Selection Framework, selected patient case examples were assessed using the framework. Cases were drawn from the UPNAT and RTLP networks and had been referred by clinicians or researchers for consideration of ASO development. These cases were intended to illustrate application of the framework in practice rather than to provide a representative or exhaustive sample.

All cases underwent initial triage using the version 1 N1C VARIANT assessment guidelines to determine variant-level amenability to ASO intervention. A subset of cases was then selected for full assessment using the complete UPNAT Target Selection Framework, incorporating disease, functional model, patient, and variant modules (**Table 1**). In keeping with the disease-agnostic design of the framework, case examples were chosen to capture diversity in affected tissue, age at assessment, and therapeutic context, including both systemically targetable and tissue-restricted diseases.

**Table 1:** Case examples selected for ASO amenability assessment using the UPNAT Target Selection Framework.

| Disease (OMIM) | Associated gene | Target tissue | Age at assessment | Assessor category |
| --- | --- | --- | --- | --- |
| Centronuclear myopathy (310400) | *MTM1* | Skeletal muscle and liver | Paediatric | Assessor 1: Academic molecular geneticist<br>Assessor 2: Academic molecular biologist<br>Assessor 3: Consultant paediatric neurologist |
| Niemann-Pick disease, type C (257220) | *NPC1* | CNS | Adolescent | Assessor 1: Academic molecular geneticist<br>Assessor 2: Consultant paediatric neurologist<br>Assessor 3: Consultant neurologist |
| Doyne honeycomb retinal dystrophy (126600) | *EFEMP1* | Retina | Adult | Assessor 1: Academic molecular geneticist, Preclinical model development expert<br>Assessor 2: Academic molecular geneticist, Preclinical model development expert<br>Assessor 3: Consultant ophthalmologist |
| BANDDOS (618476) | *CSF1R* | CNS | Paediatric | Assessor 1: Academic molecular geneticist<br>Assessor 2: Health and care professions council (HCPC) accredited clinical scientist<br>Assessor 3: Consultant paediatric neurologist<br>Assessor 4: Consultant neurologist |

All selected cases met at least one of the following criteria: (i) consideration for novel ASO development by the referring clinician; (ii) active development of a novel ASO therapy; or (iii) approval of a novel ASO for clinical trial.

## Results

Results are presented in three parts: description of the framework, calibration using approved ASO targets, and application to real-world patient case examples.

### The UPNAT Target Selection Framework

The UPNAT Target Selection Framework defines a disease-agnostic, modular system to support structured assessment of patients and targets for ASO therapies (**Figure 2**). The framework is organised into four modules, disease, functional model, patient, and variant, each addressing a distinct dimension of ASO suitability. Modules may be applied independently or in combination, allowing flexibility according to the available evidence, stage of evaluation, and clinical or research context. Together, the modules provide a common structure for evaluating biological tractability, research readiness, and patient-level considerations within a single assessment process.

**Figure 2:**
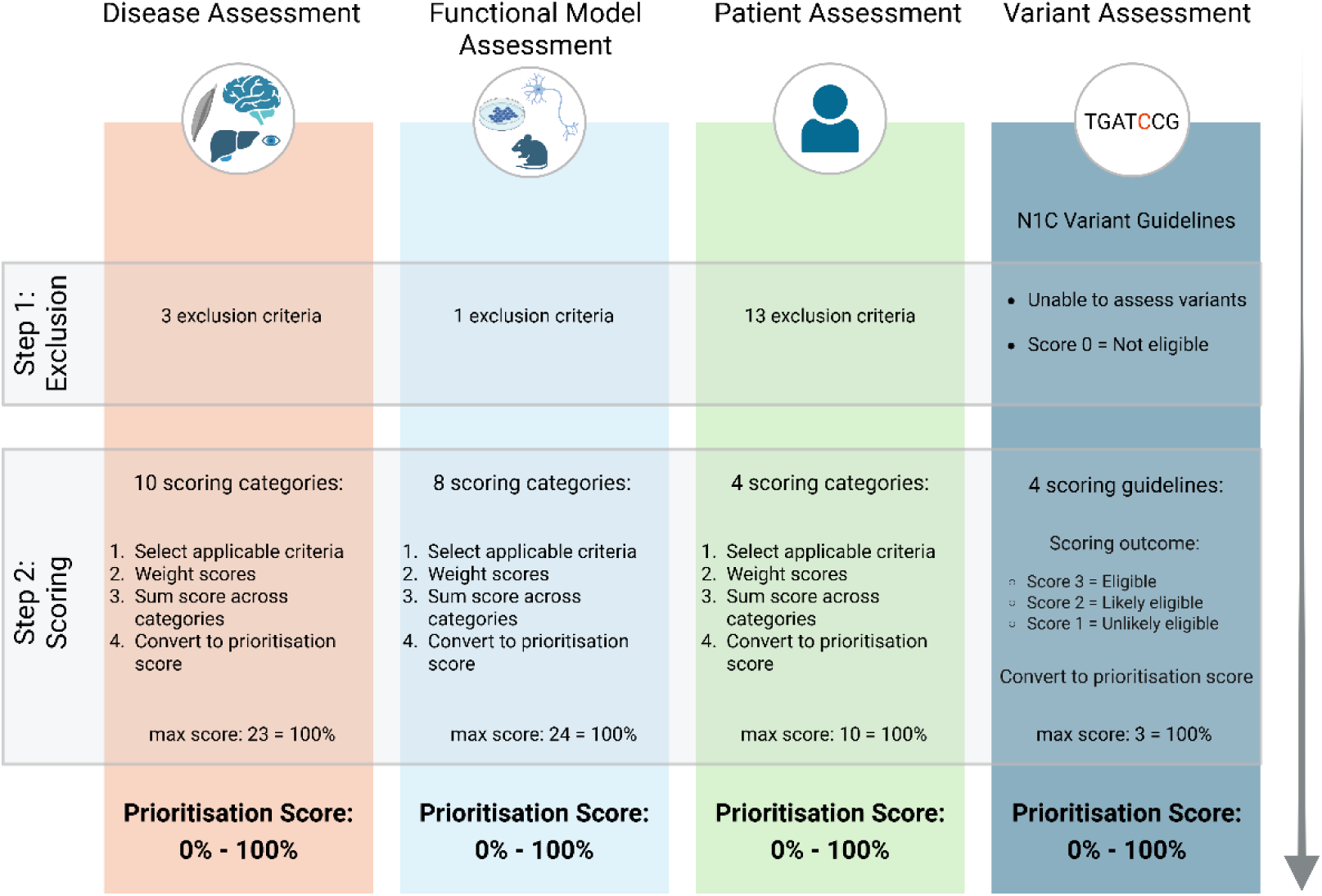
The UPNAT Target Selection Framework. The four modules address disease, functional model, patient and variant selection criteria. Each module includes Step 1 exclusion and Step 2 scoring, resulting in a prioritisation score. The variant guideline incorporates the N1C VARIANT consensus guideline (10).

Application of the framework within each module follows a two-stage process. First, predefined exclusion criteria are assessed to identify candidates unsuitable for further consideration within that domain. These criteria enable early termination where biological, technical, or clinical constraints render ASO development or treatment infeasible, conserving resources and avoiding inappropriate therapeutic expectation. Candidates not meeting exclusion criteria proceed to module-specific scoring, in which assessors select relevant predefined criteria within each category to generate a prioritisation score. Weighted categories are applied uniquely within the disease module. Scores are generated independently for each module and reflect relative prioritisation within that domain rather than absolute suitability or predicted therapeutic success. No fixed thresholds are imposed; outcomes are interpreted alongside available evidence and clinical judgement to support transparent and reproducible decision-making. Exclusion within any module precludes progression within that assessment cycle but does not prevent reassessment should new evidence, technologies, or clinical circumstances emerge. Complete module-level scoring, including selected criteria, for case examples is provided in Supplementary Table 3 to ensure full inspectability.

Modules are designed for application by assessors with domain-relevant expertise within a tiered structure, allowing assessment effort to scale with case complexity. Early exclusion criteria facilitate efficient triage, while predefined specialist-derived criteria enable consistent implementation across settings.

#### Disease module

The disease module evaluates whether a given disease represents a plausible candidate for novel ASO development. Assessment focuses on biological tractability and anticipated clinical benefit, incorporating exclusion criteria related to the expected impact of ASO intervention, the availability of existing effective therapies, and the feasibility of delivering ASOs to the affected tissue using current technologies. Diseases that pass exclusion criteria are assessed using a structured scoring framework that distinguishes technical and biological features supporting ASO development from broader prioritisation considerations. This module uniquely incorporates weighted scoring, reflecting the separation between criteria that fall within the direct scope of ASO feasibility assessment and broader considerations that address whether a target should be pursued in the absence of external national guidance (**Supplementary Table 4**). The disease module was retrospectively applied to indications with regulator-approved ASO therapies to assess concordance with established clinical translation and subsequently applied prospectively to four illustrative patient cases.

#### Functional model module

The functional model module evaluates the availability and suitability of experimental systems for assessing the efficacy of a novel ASO in the context of the target gene’s biological expression and function. Exclusion is applied when spatial or temporal gene expression is insufficiently characterised to support appropriate experimental modelling for ASO development. Candidates that proceed are evaluated based on the availability and relevance of experimental models, including evidence of gene expression across appropriate *in vitro* and *in vivo* systems, functional assays within the target cell type, and prior investigation of the gene in a therapeutic context. Together, these criteria capture the extent to which a target can be supported by existing functional knowledge, experimental infrastructure and highlight areas for improvement (**Supplementary Table 5**). The functional model module was retrospectively benchmarked against genes targeted by regulator-approved ASO therapies and prospectively applied to four illustrative patient cases.

#### Patient module

The patient module evaluates individual clinical suitability for ASO treatment using patient-specific clinical data and prognostic considerations. In contrast to the disease and functional model modules, this module is intentionally exclusion-focused, reflecting the clinical caution required when considering ASO therapies as a treatment option. Exclusion criteria address factors that may limit anticipated benefit or increase risk, while a limited set of scoring criteria is applied only after exclusion criteria are satisfied. This structure emphasises appropriateness and safety over prioritisation, recognising the ethical and clinical implications of recommending ASO treatment for individual patients (**Supplementary Table 6**). The patient module was prospectively applied to four illustrative cases in parallel with ongoing specialist evaluation for ASO therapy to assess its usability as a structured decision-support tool.

#### Variant module

The variant module assesses genetic amenability to ASO intervention by incorporating the N1C VARIANT selection guidelines (10). Variants were evaluated across multiple mechanistic strategies, including splice correction, transcript knockdown, and upregulation. Variant-level outcomes were mapped into the UPNAT framework through conversion to a unified prioritisation score, enabling integration of variant amenability with disease-, functional-, and patient-level assessments (**Figure 2**). Given that variant-level guidance is defined within the peer-reviewed N1C guidelines, this module was applied as a triage step in a UK patient cohort and integrated with disease, functional, and patient-level assessments in the illustrative cases to demonstrate end-to-end framework application.

### Framework performance of market approved ASO therapy targets

To examine how the UPNAT Target Selection Framework performs when applied to established clinical successes, targets of relevant market approved ASO therapies with well characterised backbones and safety profiles were selected (**Supplementary Table 2**). This analysis was undertaken as a calibration exercise rather than as an evaluation of predictive performance. For all regulator-approved targets, the patient assessment was redundant.

Of the eight target diseases investigated (11 ASOs in total), hereditary transthyretin amyloidosis and hereditary angioedema (**Figure 3A**) were excluded on the disease assessment due to the availability of effective alternative treatment options (21, 22). These exclusions reflect the prioritisation logic rather than a lack of biological tractability. The remaining market approved targets were not excluded on the basis of the disease and functional model assessments, indicating consistency between the framework criteria and established clinical translation.

**Figure 3:**
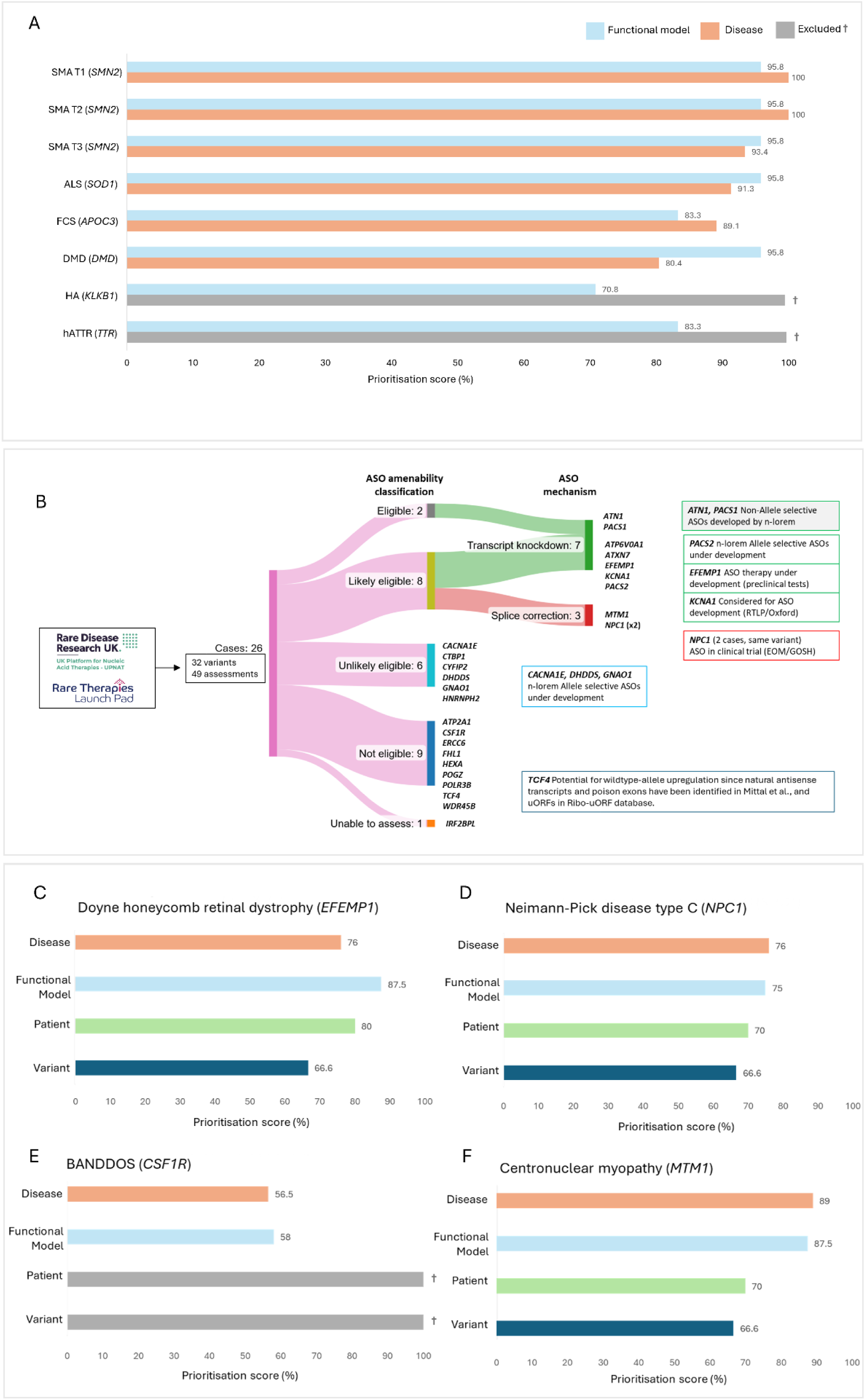
Application of the UPNAT assessment framework to targets of market approved ASOs and patient case examples referred from the UPNAT and RTLP networks. **A)** Disease and functional model assessment outcomes for targets of market-approved ASO therapies. Outcomes are displayed as prioritisation scores, representing the percentage of the maximum possible score achieved for each disease or gene. Hereditary angioedema (HA) and hereditary transthyretin amyloidosis polyneuropathy (hATTR) were excluded at the disease module stage, indicated by grey bars and † symbols. **B)** Variant assessment of the 26 individuals recruited through the UPNAT or RTLP networks. Assessments were made with N1C VARIANT guidelines (10). Nine amenable variants (‘Eligible’, or ‘Likely Eligible’) were identified for transcript knockdown or splice correction. Boxes indicate genes with ASOs either developed or under development. Grey box indicates genes assessed as ‘Eligible’. **C)** Assessment of a patient presenting with Doyne honeycomb retinal dystrophy and the causative pathogenic c.1033C>T, p.(Arg345Trp) toxic gain-of-function variant within the *EFEMP1* gene. **D)** Assessment of a patient presenting with Neiman-Pick disease type C and the causative pathogenic c.1554-1009G>A splice disrupting variant within the *NPC1* gene. **E)** Assessment of a patient presenting with BANDDOS and the pathogenic splice disrupting variant c.592+5 A>G in trans with a pathogenic nonsense variant within the *CSF1R* gene. Both variants were assessed as ‘Not Eligible’. Exclusion depicted by a grey bar and † symbol. **F)** Assessment of a patient presenting with centronuclear myopathy and the causative pathogenic target c.342+575C>G splice disrupting variant within the *MTM1* gene. ALS, amyotrophic lateral sclerosis; FCS, familial chylomicronemia syndrome; DMD, Duchenne muscular dystrophy.

Spinal muscular atrophy (SMA) was assessed for each clinical subtype recommended for treatment with nusinersen by NICE (**Figure 3A**). Variation between the SMA clinical subtypes was observed within the disease module assessment, reflecting the difference in disease severity. For all market approved ASO therapies, variant-level scoring was not applicable, as therapeutic strategies were either variant agnostic or used mechanisms not compatible with variant-specific assessment within the framework.

### Application of the UPNAT Framework to a real-world patient cohort

To demonstrate the real-world usability of the UPNAT Target Selection Framework, patient case examples that had been identified by clinicians or researchers as potential candidates for ASO evaluation were assessed using the framework. In this implementation, assessment was initiated at the variant level, reflecting current data availability and the existence of established variant-focused guidance through the version 1 N1C VARIANT selection guidelines. Consistent with its modular design, the framework does not depend on variant-first entry, and as assessment processes evolve, alternative entry points may become more appropriate for large-scale deployment.

Following initial variant-level triage, 10 of 26 cases were classified as ‘Eligible’ or ‘Likely Eligible’, with the remainder excluded at this stage. Among these, three cases were classified as amenable to splice correction and seven to transcript knockdown (**Supplementary Table 7** and **Figure 3B**). No cases met formal criteria for upregulation.

#### Splice correction

Eight splice-disrupting variants were assessed for splice correction. Two variants were classified as ‘Likely Eligible’. The first was an intronic NM_000271.5:c.1554-1009G>A *NPC1* variant identified *in trans* with missense variants in two unrelated cases within the cohort. Aberrant splicing was confirmed by RNA sequencing in both patients, and the variant met eligibility criteria for splice correction based on its deep intronic location and predicted preservation of canonical splicing. A second variant was a deep intronic NM_000252.3:c.342+575C>G change within the *MTM1* gene. Functional studies confirming intronic retention (or pseudo exon inclusion), combined with the deep intronic position of the variant, supported its eligibility for splice-corrective therapy. All remaining splice variants resided within canonical splice sites and were therefore not considered eligible for splice-correcting ASOs under N1C criteria. Of the 24 variants considered for exon skipping approaches, a single variant in *PACS1* was considered ‘Unlikely Eligible’ and the remaining were ‘Not Eligible’, primarily due to predicted out-of-frame exon skipping, disruption of functional domains, or localisation within single-exon genes.

#### Transcript knockdown

Fourteen gain-of-function variants were evaluated for transcript knockdown. Seven variants were classified as ‘Eligible’ or ‘Likely Eligible’ in genes without evidence of haploinsufficiency in the target tissue. These included the NM_001039348.3:c.1033C>T(p.Arg345Trp) variant within the *EFEMP1* gene, which has since been targeted by a novel gene-silencing ASO, with preclinical data supporting variant eligibility (23). Six variants were classified as ‘Unlikely Eligible’, and one as ‘Not Eligible’, due to established haploinsufficiency or intolerance to reduction in the corresponding genes.

#### WT transcript upregulation

Three variants were considered for wild-type transcript upregulation. No variants were considered ‘Eligible’ under N1C guidelines. The frameshift variant within *IRF2BPL* was deemed unable to assess due to an unknown pathogenic mechanism and the remaining two variants were deemed ‘Not Eligible’.

From the cohort of 26 individuals assessed for variant guidelines, four real-world case examples were selected for detailed assessment to demonstrate how the full UPNAT framework operates across distinct decision scenarios (**Figure 3C-F**).

### Case Example 1: Prioritisation supported by cross-module concordance

#### Doyne honeycomb retinal dystrophy (*EFEMP1)*

Doyne honeycomb retinal dystrophy (DHRD) is a progressive inherited retinal dystrophy caused by an autosomal dominant toxic gain-of-function variant in *EFEMP1*. A patient with a confirmed *EFEMP1* (NM_001039348.3):c.1033C>T(p.Arg345Trp) variant was assessed following continued progression of visual impairment despite standard management (**Figure 3C**). Disease and functional model modules generated scores in line with those reported for clinically translated market approved targets (**Figure 3A**), supporting prioritisation of DHRD and *EFEMP1* as targets.

#### Disease module

DHRD did not meet any exclusion criteria and generated a disease score of 76%. This reflected established targetability of retinal tissue (24, 25), a well-defined toxic gain-of-function disease mechanism (26–30), a comprehensive natural history study (31), and the availability of clinically meaningful outcome measures (26, 32), in the absence of effective disease-modifying therapies.

#### Functional model module

*EFEMP1* generated a prioritisation score of 87.5%, reflecting established expression in the retina and the availability of relevant in vitro and in vivo models and validated functional assays to support preclinical ASO development (28–30, 33–39).

#### Patient module

The patient did not meet any exclusion criteria and scored 80%, reflecting the presence of measurable clinical endpoints and an overall favourable anticipated benefit–risk profile for ASO intervention.

#### Variant module

The *EFEMP1* c.1033C>T (p.Arg345Trp) variant was classified as ‘Likely Eligible’ for transcript knockdown based on its tissue-specific toxic gain-of-function mechanism and predicted tolerance of partial transcript reduction. Haploinsufficiency assessment was complicated by available data suggesting that *EFEMP1* haploinsufficiency underlies a connective tissue disorder (40). However, retinal tissue was not implicated and, whilst *EFEMP1* is expressed ubiquitously, ASOs are delivered to the retina via intravitreal injection. The absence of retinal tissue involvement in reported haploinsufficiency and autosomal recessive loss-of-function cases supports continued knockdown targeting.

### Case Example 2: Prioritisation with translational constraints

#### Niemann–Pick disease type C (*NPC1*)

A splice-switching ASO targeting the intronic *NPC1* variant c.1554-1009G>A has been approved for the UK’s first individualised ASO clinical trial at Great Ormond Street Hospital (41, 42). Niemann–Pick disease type C (NPC) is an autosomal recessive neurodegenerative disorder caused by biallelic loss-of-function variants in *NPC1* (43). The c.1554-1009G>A variant introduces a pseudo-exon and has been confirmed as pathogenic through RNA sequencing and functional assays (44). A patient with childhood-onset disease and progressive neurological decline was assessed (**Figure 3D**). The disease and functional model modules generated scores comparable to those observed for regulator-approved ASO targets (**Figure 3A**), supporting prioritisation of *NPC1* as a therapeutic target, while framework criteria relating to disease resolution and extent of disease targeting indicate potential translational constraints consistent with partial therapeutic rescue.

#### Disease module

NPC generated a disease score of 76%, supported by central nervous system (CNS) targetability, a well-characterised disease mechanism (45), established longitudinal natural history studies (43, 46–48) and clinical outcome measures. The presence of miglustat as an approved disease-modifying therapy was incorporated into the scoring framework, with weighting informed by specialist clinical assessment of its modest efficacy.

#### Functional model module

*NPC1* generated a functional model score of 75%, reflecting the availability of established cellular assays (49) and animal models (50), alongside limitations including a humanised model optimised for ASO evaluation.

#### Patient module

The patient did not meet exclusion criteria and generated a patient prioritisation score of 70%, reflecting disease stage compatible with intervention and acceptable anticipated risk–benefit balance.

#### Variant module

The deep intronic c.1554-1009G>A variant was classified as ‘Likely Eligible’ for splice correction under the N1C VARIANT guidelines, translating to a prioritisation score of 66.6%.

### Case Example 3: Exclusion driven by variant- and patient-level criteria

#### Brain abnormalities, neurodegeneration, and dysosteosclerosis (*CSF1R*)

This case illustrates exclusion at the patient and variant level despite biological tractability. A child with brain abnormalities, neurodegeneration, and dysosteosclerosis (BANDDOS) caused by compound heterozygous loss-of-function variants in *CSF1R* was considered for ASO intervention targeting only the CNS disease manifestations (**Figure 3E**). BANDDOS is a severe recessive disorder characterised by progressive neurodegeneration and skeletal abnormalities, with no disease-modifying therapies currently available.

#### Disease module

BANDDOS generated a disease score of 56.5%, supported by disease severity, absence of alternative treatments, CNS targetability, and a defined disease mechanism (51). Limitations included sparse natural history data and lack of established clinical endpoints.

#### Functional model module

The functional model score was 58%, reflecting availability of basic *in vitro* systems and prior therapeutic investigation of *CSF1R* (51), alongside limitations in model relevance for ASO development.

#### Patient module

The patient met a Step 1 exclusion criterion, reflecting an advanced and rapidly progressive disease stage and an unfavourable anticipated risk–benefit balance for ASO intervention at the time of assessment.

#### Variant module

Molecular investigation confirmed one nonsense and one splice-altering allele (52). Splice correction was not supported for the pathogenic variants, and although upregulation strategies were considered, these were not pursued given the association of *CSF1R* haploinsufficiency with a milder adult-onset disorder and the current immaturity of upregulation approaches.

### Case Example 4: Conditional progression limited by translational constraints

#### Centronuclear myopathy (*MTM1*)

This case illustrates high biological and variant-level readiness tempered by technical constraints related to delivery. Centronuclear myopathy is a severe X-linked recessive disorder caused by loss-of-function variants in *MTM1*, characterised by profound muscle weakness and early morbidity (53). A patient with a deep intronic hemizygous *MTM1* NM_000252.3:c.342+575C>G variant was assessed (**Figure 3F**), with a splice-correction ASO strategy under exploration. While disease and functional module scores suggest biological and technical feasibility comparable to regulator-approved ASO targets (**Figure 3A**), translational challenges related to efficient delivery to skeletal muscles, together with specialist clinical caution, temper prioritisation in centronuclear myopathy.

#### Disease module

Centronuclear myopathy generated a disease score of 89%, reflecting a well-defined disease mechanism, comprehensive natural history data, established clinical endpoints, and absence of effective therapies (53–55). The disease affects skeletal muscle and liver, both of which have evidence of ASO targetability, with technical limitations in clinical efficacy acknowledged.

#### Functional model module

*MTM1* generated a functional model score of 87.5%, supported by extensive *in vitro* and *in vivo* models, well-characterised gene function, and established functional assays relevant to ASO development.

#### Patient module

The patient did not meet exclusion criteria and generated a patient prioritisation score of 70%, reflecting an acceptable anticipated risk–benefit balance.

#### Variant module

The deep intronic c.342+575C>G variant met criteria for ‘Likely Eligible’ splice correction under the N1C VARIANT guidelines. Translation to therapy would, however, necessitate effective ASO delivery to both skeletal muscle and liver, with recognised technical limitations in achieving efficient skeletal muscle delivery noted when considering the ASO chemistries currently approved for neuromuscular disorders.

Together, these case examples illustrate how different constraints dominate decision-making at different stages of assessment. In some cases, strong alignment across disease, functional, patient, and variant modules support prioritisation, while in others, patient-level considerations or technical feasibility shape progression or exclusion despite biological tractability. Rather than collapsing these factors into a single outcome, the modular structure of the UPNAT framework makes the basis of each decision explicit, supporting transparent, reproducible, and revisable prioritisation as evidence, technologies, and therapeutic capabilities evolve.

## Discussion

The UPNAT Target Selection Framework was developed to address a critical gap in the translational pathway for ASO therapies: the absence of structured, transparent, and scalable approaches to prioritising patients and targets (diseases, genes, or variants) within a publicly funded healthcare system. While advances in genomic diagnostics and NATs have expanded the range of potentially actionable rare diseases, decision-making around ASO development has largely remained informal, fragmented, and dependent on local expertise. Such decisions potentially vary substantially between institutions and are rarely documented in a form that permits comparison, audit, or re-evaluation. UPNAT formalises this decision space without reducing it to a single predictive metric, providing instead a modular framework that makes the basis for progression, deferral, or exclusion explicit and reviewable.

A defining feature of the framework is its separation of biological feasibility from clinical and system-level prioritisation. By deconstructing assessment into disease, functional model, patient, and variant modules, this framework enables structured evaluation across dimensions that are often conflated in practice. This modularity reflects the reality that suitability for ASO intervention is multi-dimensional: a variant may be mechanistically amenable while disease context, model readiness, or patient-level considerations limit feasibility or justify caution. Importantly, module scores are not intended to function as binary decision thresholds or predictors of therapeutic success. Rather, they support relative prioritisation within domains, highlight sources of uncertainty, and document the rationale underpinning decisions. This design recognises that ASO development involves iterative judgement under uncertainty, rather than deterministic selection based on any single criterion.

Calibration of the framework against targets of market-approved ASO therapies demonstrated that this framework behaves in a manner that is consistent with established translational outcomes. Approved targets scored highly on biological tractability and functional model readiness, while exclusions arising from the disease module reflected the availability of effective alternative treatments rather than a lack of mechanistic suitability. These findings should not be interpreted as validation of predictive performance. Instead, they indicate that the framework encodes prioritisation logic that aligns with real-world decision-making, providing a structured representation of assessments that are currently made implicitly and inconsistently. In doing so, UPNAT Target Selection Framework enables greater consistency, scrutiny, and reproducibility in clinical decision-making.

Application of the framework to real-world patient case examples further illustrates its intended use. Entry into assessment was initiated at the variant level in this study, reflecting current data availability and the existence of established variant-focused guidance. However, the framework is explicitly modular and does not depend on a variant-first workflow. As assessment processes evolve, including increasing automation and broader disease-level screening, alternative entry points may become more appropriate, particularly for large-scale deployment.

The four detailed case examples were selected to illustrate distinct decision scenarios rather than to maximise apparent success. Together, they demonstrate how the framework supports prioritisation when biological, experimental, and patient-level factors align; principled exclusion based on variant- and patient-level considerations despite biological tractability; and recognition of technical constraints that may limit amenability to ASO therapy even in the presence of strong scores across multiple modules. Importantly, exclusion within the framework does not imply definitive or permanent ineligibility. Instead, it documents the specific constraints operating at the time of assessment, enabling re-evaluation as evidence, technologies, or clinical circumstances change.

Application of the framework also highlighted limitations inherent to retrospective assessment and to the current evidentiary landscape for ASO development. Publicly available data are heavily skewed towards successful or advanced ASO programmes, while detailed information on targets that were deprioritised or failed to progress is rarely accessible. This introduces unavoidable bias and constrains systematic refinement of prioritisation criteria, underscoring the need for greater transparency and data sharing to support responsible expansion of NATs.

Variant-level triage proved to be a pragmatic and low-cost entry point for assessment, reflecting current clinical data availability and the maturity of existing variant-focused guidance. However, its application also exposed important gaps. Deep intronic variants were frequently difficult to evaluate in the absence of functional validation and remain under-represented in routine clinical reporting, as most diagnostic pipelines prioritise exonic and canonical splice-site regions. Similarly, while allele-specific transcript knockdown emerged as a potentially actionable strategy in several cases, its feasibility depended on confidence in gene dosage tolerance. In practice, haploinsufficiency assessment was often complicated by discordant bioinformatic metrics, incomplete population reference data, and tissue-specific effects, reinforcing the need to interpret variant amenability in the context of disease biology and affected tissue. These considerations were evident across case examples, including DHRD, centronuclear myopathy and BANDDOS, where biological tractability alone was insufficient to determine strategy selection without careful evaluation of tissue specificity, dual targeting requirements, underlying disease mechanism, and anticipated risk–benefit. Importantly, such uncertainties do not represent failures of the framework but rather reflect current limits of knowledge, which the modular structure is designed to surface explicitly rather than obscure through aggregate scoring. In addition, efficient and cell-specific *in vivo* delivery of ASOs remains a major translational challenge. Advances in ASO chemistries, conjugates and delivery systems are likely to expand tissue targeting capabilities and improve safety and efficacy, making this an area of ongoing evolution that may alter feasibility assessments over time.

The clinical relevance of this structured approach is illustrated by the NPC case, in which framework assessment was conducted alongside specialist-led evaluation preceding an individualised ASO clinical trial in the UK. In this context, use of a master protocol provides a scalable model for future enrolment as evidence evolves, consistent with the framework’s emphasis on reproducibility, iterative re-evaluation, and responsible expansion of access rather than one-off decision-making.

More broadly, the UPNAT Target Selection Framework provides a structured basis for both governance and prioritisation. By standardising how decisions are organised and recorded, it enables scrutiny, comparison, and iterative refinement across cases and institutions. Disease biology and functional model readiness are intended for episodic specialist assessment, with gene- and disease-level determinations reusable across centres, whereas patient-level evaluation is designed for iterative local application under appropriate oversight. Although variant assessment may be undertaken locally, coordinated genomic systems with automated annotation capacity may benefit from more centralised approaches to improve efficiency and scalability. Evaluations should be regarded as dynamic and subject to revision as new clinical, molecular, and therapeutic data emerge. Where appropriate, non-confidential components could be shared transparently, recognising that many domains reflect gene- or disease-level considerations rather than individual patient characteristics and that such sharing may support cumulative learning across therapeutic modalities.

This structured approach is particularly relevant for healthcare systems seeking equitable and accountable delivery of advanced NATs. While developed using ASOs as a pilot modality, the underlying principles are not platform-specific and are likely to extend to other programmable nucleic acid–based therapeutics. As these modalities expand across rare and more common diseases, scalable and transparent decision architectures will be essential to balance innovation with responsible clinical implementation. The modular governance model presented here therefore offers a transferable framework to support reproducible prioritisation and equitable access to advanced genetic therapies.

Several limitations should be acknowledged. The framework does not aim to predict clinical efficacy, nor does it resolve all uncertainties inherent in ASO development. Its performance is contingent on the quality of available evidence, and some modules may be more informative than others depending on disease context. In addition, the case examples presented here reflect a curated cohort with prior indication of ASO amenability and should not be interpreted as representative of all rare disease referrals. Ongoing application and refinement, including prospective use within specialised services, will be essential to assess usability, consistency, and impact on translational decision-making. Further validation against regulator-approved and clinically translated ASO targets will refine interpretation of prioritisation scores, particularly as evaluation expands across diverse ASO chemistries and the growing number of n-of-1 and small-cohort therapies.

In summary, the UPNAT Target Selection Framework provides a structured, transparent approach to prioritising ASO development within a publicly funded healthcare system. By making decision logic explicit and modular, the framework supports equitable and reproducible assessment while accommodating uncertainty and evolution in therapeutic capability. As precision NATs continue to expand, the absence of such framework risks amplifying inconsistency rather than access. UPNAT offers a practical model for aligning translational opportunities with responsible, consistent, and patient-centred decision-making.

## Acknowledgements

The authors would like to acknowledge the initiation of this platform, funding, and project management support from UPNAT. UPNAT is led by Prof Haiyan Zhou as the Principal Investigator/Director, Prof Francesco Muntoni as Deputy Director, and is managed by Ms Catherine Ryan. UPNAT is hosted at University College London. UPNAT is funded by the National Institute for Health and Care Research (NIHR) & the Medical Research Council (MR/Y008405/1 to H.Z.). NATA is funded by MRC Programme Grant MC_PC_20061.

This framework has been supported by NIHR Biomedical Research Centre (BRC) infrastructure including NIHR Great Ormond Street Hospital BRC, NIHR BRC at Moorfields Eye Hospital and UCL Institute of Ophthalmology, NIHR UCLH BRC, NIHR Cambridge BRC and NIHR Oxford BRC (J.C.T., C.C. and A.G.L.D.). We thank Genetic Alliance UK and Unique for providing patient advocacy oversight and engagement through the framework design and development process. We also acknowledge EveryONE Medicines and N=1 Collaborative for their critical appraisal of the framework. We thank Jillian Hastings and David Jones for their critical appraisal of this manuscript on behalf of RDR UK. We thank Genomics England for their support of the UPNAT program and acknowledge the contributions of A.L.T. and N.E. for their valuable expertise and time dedicated to this work, supported by Genomics England.

A.E.D. is funded by a UKRI Future Leader Fellowship MR/S031820/1 and MR/Y019911/1.

This work was supported by the Medical Research Council Centre of Research Excellence (CoRE) in Therapeutic Genomics (grant number MR/Z504725/1 to S.J.S. and J.C.T.) and the Health Data Research UK QQ2 Molecules to Health Records Driver Programme (to SJS).

Figure 1D and Figure 2 generated using Biorender.com.

## Author contributions

H.Z., M.R., and A.E.D. conceived the Target Selection Framework project and secured funding for this study.

H.Z., F.M., M.R., A.E.D., C.R., C.R.y., and N.L. formed the UPNAT committee and provided strategic oversight of UPNAT platform development.

H.Z., M.R. and A.E.D. conceived the framework. M.R. and A.E.D led the overall framework development, with foundational supervision from H.Z.. The disease module was led by E.C. and H.T.; the functional model module by J.v.d.S. and S.J.S.; the patient module by C.R. and M.A.K.; and the variant module by

J.C.T. and A.L.T., with N.E. and C.C. contributing to variant module development. F.M. provided expert advisory input to framework development. E.F.W. coordinated framework integration and cross-module synthesis.

E.F.W. coordinated and implemented the framework across market-approved targets and case example evaluations.

Framework calibration using approved ASO therapies was performed by E.F.W., K.-A.M., M.R., C.R., F.M. and H.Z.

Variant-level assessments were performed by C.C., N.E., J.C.T. and E.F.W..

Clinical case identification, evaluation and specialist clinical input were provided by F.M., U.K., H.S., A.G.L.D., L.W., H.G.L., D.S.L., R.R., M.M., T.A.C.G., M.R., and P.G..

Disease and functional model module assessments for patient case examples were supported by P.G., M.R., D.S.L., K.-A.M., E.F.W., J.v.d.S., A.E.D, T.A.C.G., M.M., and S.A..

E.F.W. drafted the original manuscript. M.R. and A.E.D. provided substantial input to manuscript development and revisions leading to the final version. C.C., N.E. and J.C.T. provided the variant assessment text, figure and supplementary table. With exception to the variant figure and supplementary table, all figures and tables designed and generated by E.F.W., with input from M.R, A.E.D. and K.-A.M.. K.-A.M., C.R., C.R.y., F.M. and H.Z. provided important feedback and revisions during drafting.

All authors contributed to manuscript review, critical revision and approval of the final version.

## Ethics statement

This study describes the development of a decision-support framework and its application to previously reported or clinically referred cases. No prospective or interventional research was conducted. De-identified clinical and genetic data were used in accordance with existing governance frameworks and relevant institutional and national guidelines (IRAS: 141100, 95005, and 204280).

## Data availability

Data supporting the findings of this study are included within the article and its Supplementary Information. Additional information may be available from the corresponding authors upon reasonable request, subject to applicable data governance and patient confidentiality restrictions.

## Code availability

No custom code was developed or used in this study.

## References

1. The Lancet Global H. The landscape for rare diseases in 2024. Lancet Glob Health. 2024;12(3):e341.

2. Finkel RS, Mercuri E, Darras BT, Connolly AM, Kuntz NL, Kirschner J, et al. Nusinersen versus Sham Control in Infantile-Onset Spinal Muscular Atrophy. N Engl J Med. 2017;377(18):1723–32.

3. Raal FJ, Santos RD, Blom DJ, Marais AD, Charng MJ, Cromwell WC, et al. Mipomersen, an apolipoprotein B synthesis inhibitor, for lowering of LDL cholesterol concentrations in patients with homozygous familial hypercholesterolaemia: a randomised, double-blind, placebo-controlled trial. Lancet. 2010;375(9719):998–1006.

4. Wheeler TM, Leger AJ, Pandey SK, MacLeod AR, Nakamori M, Cheng SH, et al. Targeting nuclear RNA for in vivo correction of myotonic dystrophy. Nature. 2012;488(7409):111–5.

5. Modarresi F, Faghihi MA, Lopez-Toledano MA, Fatemi RP, Magistri M, Brothers SP, et al. Inhibition of natural antisense transcripts in vivo results in gene-specific transcriptional upregulation. Nat Biotechnol. 2012;30(5):453–9.

6. Roberts TC, Langer R, Wood MJA. Advances in oligonucleotide drug delivery. Nat Rev Drug Discov. 2020;19(10):673–94.

7. Zarouchlioti C, Sanchez-Pintado B, Hafford Tear NJ, Klein P, Liskova P, Dulla K, et al. Antisense Therapy for a Common Corneal Dystrophy Ameliorates TCF4 Repeat Expansion-Mediated Toxicity. Am J Hum Genet. 2018;102(4):528–39.

8. Kim J, Hu C, Moufawad El Achkar C, Black LE, Douville J, Larson A, et al. Patient-Customized Oligonucleotide Therapy for a Rare Genetic Disease. N Engl J Med. 2019;381(17):1644–52.

9. Chopra M, Modi ME, Dies KA, Chamberlin NL, Buttermore ED, Brewster SJ, et al. GENE TARGET: A framework for evaluating Mendelian neurodevelopmental disorders for gene therapy. Mol Ther Methods Clin Dev. 2022;27:32–46.

10. Cheerie D, Meserve MM, Beijer D, Kaiwar C, Newton L, Taylor Tavares AL, et al. Consensus guidelines for assessing eligibility of pathogenic DNA variants for antisense oligonucleotide treatments. Am J Hum Genet. 2025;112(5):975–83.

11. The Medicines for Human Use (Clinical Trials) (Amendment) Regulations 2025.

12. Agency MaHpR. Rare therapies and UK regulatory considerations.

13. Belgrad J, McConnell E, Leonard S, Nolen N, Lauffer MC, Watts JK, et al. The N=1 Collaborative: advancing customized nucleic acid therapies through collaboration and data sharing. Nucleic Acids Res. 2025;53(8).

14. Synofzik M, van Roon-Mom WMC, Marckmann G, van Duyvenvoorde HA, Graessner H, Schule R, et al. Preparing n-of-1 Antisense Oligonucleotide Treatments for Rare Neurological Diseases in Europe: Genetic, Regulatory, and Ethical Perspectives. Nucleic Acid Ther. 2022;32(2):83–94.

15. Aartsma-Rus A. OTS Rare Disease N-of-1+ Workshop Briefing Document. Oligonucleotide Therapeutics Society 2020.

16. Kim J, Woo S, de Gusmao CM, Zhao B, Chin DH, DiDonato RL, et al. A framework for individualized splice-switching oligonucleotide therapy. Nature. 2023;619(7971):828–36.

17. Zardetto B, Lauffer MC, van Roon-Mom W, Aartsma-Rus A, On Behalf Of The NC. Practical Recommendations for the Selection of Patients for Individualized Splice-Switching ASO-Based Treatments. Hum Mutat. 2024;2024:9920230.

18. Wijnant KN, Nadif Kasri N, Vissers L. Systematic analysis of genetic and phenotypic characteristics reveals antisense oligonucleotide therapy potential for one-third of neurodevelopmental disorders. Genome Med. 2025;17(1):59.

19. Schule R, Graessner H, Aartsma-Rus A, van Roon-Mom WMC, Collaborative N, Mutation 1 Medicine C, et al. Tailored antisense oligonucleotides for ultrarare CNS diseases: An experience-based best practice framework for individual patient evaluation. Mol Ther Nucleic Acids. 2025;36(3):102615.

20. O’Connor DJ, Moss P, Wood M, Murphy M, Parker M, Blackwood N, et al. The Rare Therapies Launchpad: a pilot program for individualized medicines in the UK. Nat Med. 2025;31(5):1379–80.

21. Adams D, Gonzalez-Duarte A, O’Riordan WD, Yang CC, Ueda M, Kristen AV, et al. Patisiran, an RNAi Therapeutic, for Hereditary Transthyretin Amyloidosis. N Engl J Med. 2018;379(1):11–21.

22. Betschel SD, Banerji A, Busse PJ, Cohn DM, Magerl M. Hereditary Angioedema: A Review of the Current and Evolving Treatment Landscape. J Allergy Clin Immunol Pract. 2023;11(8):2315–25.

23. Farah. O. Rezek BS-P, Emily R. Eden, Nancy Aychoua, Andrew R. Webster, Amanda-Jayne F. Carr, Michel Michaelides, View ORCID ProfileMichael E. Cheetham, Jacqueline van der Spuy. Antisense oligonucleotide allele-specific targeting of EFEMP1 in a patient-derived model of Doyne honeycomb retinal dystrophy. 2025.

24. Geary RS, Henry SP, Grillone LR. Fomivirsen: clinical pharmacology and potential drug interactions. Clin Pharmacokinet. 2002;41(4):255–60.

25. Li W, Li Y, Zhou Y, Liu Y, Liu H, Wei X, et al. Exon Skipping Therapy Restores Ciliary Function in USH2A-Related Retinal Degeneration. Invest Ophthalmol Vis Sci. 2025;66(12):46.

26. Michaelides M, Jenkins SA, Brantley MA, Jr., Andrews RM, Waseem N, Luong V, et al. Maculopathy due to the R345W substitution in fibulin-3: distinct clinical features, disease variability, and extent of retinal dysfunction. Invest Ophthalmol Vis Sci. 2006;47(7):3085–97.

27. Garland DL, Fernandez-Godino R, Kaur I, Speicher KD, Harnly JM, Lambris JD, et al. Mouse genetics and proteomic analyses demonstrate a critical role for complement in a model of DHRD/ML, an inherited macular degeneration. Hum Mol Genet. 2014;23(1):52–68.

28. Sohn EH, Wang K, Thompson S, Riker MJ, Hoffmann JM, Stone EM, et al. Comparison of drusen and modifying genes in autosomal dominant radial drusen and age-related macular degeneration. Retina. 2015;35(1):48–57.

29. Marmorstein LY, Munier FL, Arsenijevic Y, Schorderet DF, McLaughlin PJ, Chung D, et al. Aberrant accumulation of EFEMP1 underlies drusen formation in Malattia Leventinese and age-related macular degeneration. Proc Natl Acad Sci U S A. 2002;99(20):13067–72.

30. Klenotic PA, Munier FL, Marmorstein LY, Anand-Apte B. Tissue inhibitor of metalloproteinases-3 (TIMP-3) is a binding partner of epithelial growth factor-containing fibulin-like extracellular matrix protein 1 (EFEMP1). Implications for macular degenerations. J Biol Chem. 2004;279(29):30469–73.

31. de Guimaraes TAC, Kalitzeos A, Mahroo OA, van der Spuy J, Webster AR, Michaelides M. A Long-Term Retrospective Natural History Study of EFEMP1-Associated Autosomal Dominant Drusen. Invest Ophthalmol Vis Sci. 2024;65(6):31.

32. Georgiou M, Robson AG, Fujinami K, de Guimaraes TAC, Fujinami-Yokokawa Y, Daich Varela M, et al. Phenotyping and genotyping inherited retinal diseases: Molecular genetics, clinical and imaging features, and therapeutics of macular dystrophies, cone and cone-rod dystrophies, rod-cone dystrophies, Leber congenital amaurosis, and cone dysfunction syndromes. Prog Retin Eye Res. 2024;100:101244.

33. Daniel S, Hulleman JD. Exploring ocular fibulin-3 (EFEMP1): Anatomical, age-related, and species perspectives. Biochim Biophys Acta Mol Basis Dis. 2024;1870(6):167239.

34. Galloway CA, Dalvi S, Hung SSC, MacDonald LA, Latchney LR, Wong RCB, et al. Drusen in patient-derived hiPSC-RPE models of macular dystrophies. Proc Natl Acad Sci U S A. 2017;114(39):E8214–E23.

35. Tsai YT, Li Y, Ryu J, Su PY, Cheng CH, Wu WH, et al. Impaired cholesterol efflux in retinal pigment epithelium of individuals with juvenile macular degeneration. Am J Hum Genet. 2021;108(5):903–18.

36. Crowley MA, Garland DL, Sellner H, Banks A, Fan L, Rejtar T, et al. Complement factor B is critical for sub-RPE deposit accumulation in a model of Doyne honeycomb retinal dystrophy with features of age-related macular degeneration. Hum Mol Genet. 2023;32(2):204–17.

37. Hulleman JD, Kaushal S, Balch WE, Kelly JW. Compromised mutant EFEMP1 secretion associated with macular dystrophy remedied by proteostasis network alteration. Mol Biol Cell. 2011;22(24):4765–75.

38. Fernandez-Godino R, Bujakowska KM, Pierce EA. Changes in extracellular matrix cause RPE cells to make basal deposits and activate the alternative complement pathway. Hum Mol Genet. 2018;27(1):147–59.

39. Zhou M, Weber SR, Zhao Y, Chen H, Barber AJ, Grillo SL, et al. Expression of R345W-Fibulin-3 Induces Epithelial-Mesenchymal Transition in Retinal Pigment Epithelial Cells. Front Cell Dev Biol. 2020;8:469.

40. Forghani I, Lang SH, Rodier MJ, Bivona SA, Undiagnosed Diseases N, Morales AA, et al. EFEMP1 haploinsufficiency causes a Marfan-like hereditary connective tissue disorder. Am J Med Genet A. 2024;194(6):e63556.

41. Treatment of a teenager with an ultra-rare condition is a medical milestone [press release]. The Economist 2026.

42. Science DoHS. England Rare Diseases Action Plan 2026: main report. In: Care HaS, editor. 2026.

43. Guatibonza Moreno P, Pardo LM, Pereira C, Schroeder S, Vagiri D, Almeida LS, et al. At a glance: the largest Niemann-Pick type C1 cohort with 602 patients diagnosed over 15 years. Eur J Hum Genet. 2023;31(10):1108–16.

44. Rodriguez-Pascau L, Coll MJ, Vilageliu L, Grinberg D. Antisense oligonucleotide treatment for a pseudoexon-generating mutation in the NPC1 gene causing Niemann-Pick type C disease. Hum Mutat. 2009;30(11):E993–E1001.

45. Carstea ED, Morris JA, Coleman KG, Loftus SK, Zhang D, Cummings C, et al. Niemann-Pick C1 disease gene: homology to mediators of cholesterol homeostasis. Science. 1997;277(5323):228–31.

46. Solomon BI, Munoz AM, Sinaii N, Mohamed H, Farhat NM, Alexander D, et al. Swallowing characterization of adult-onset Niemann-Pick, type C1 patients. Orphanet J Rare Dis. 2024;19(1):231.

47. Imrie J, Heptinstall L, Knight S, Strong K. Observational cohort study of the natural history of Niemann-Pick disease type C in the UK: a 5-year update from the UK clinical database. BMC Neurol. 2015;15:257.

48. Imrie J, Dasgupta S, Besley GT, Harris C, Heptinstall L, Knight S, et al. The natural history of Niemann-Pick disease type C in the UK. J Inherit Metab Dis. 2007;30(1):51–9.

49. Vanier MT, Latour P. Laboratory diagnosis of Niemann-Pick disease type C: the filipin staining test. Methods Cell Biol. 2015;126:357–75.

50. Pallottini V, Pfrieger FW. Understanding and Treating Niemann-Pick Type C Disease: Models Matter. Int J Mol Sci. 2020;21(23).

51. Peng L, Fang Z, Zhang W, Rao GW, Zheng Q. Recent advances in colony stimulating factor-1 receptor (CSF1R) inhibitors. Biochem Pharmacol. 2025;242(Pt 1):117187.

52. Kylie Montgomery HM, Claire Anderson, Charles Wade, Emil K Gustavsson, David S Lynch, Louise C Wilson, James Davison, Emma Wakeling, Karin Tuschl, Henry Houlden, Emma Clement, Phillipa Mills, Mina Ryten. RAPID: A Targeted Long-Read RNA Workflow for Functional Resolution of Splicing Variants in Rare Disease 2026.

53. Amburgey K, Tsuchiya E, de Chastonay S, Glueck M, Alverez R, Nguyen CT, et al. A natural history study of X-linked myotubular myopathy. Neurology. 2017;89(13):1355–64.

54. Jungbluth H, Gautel M. Pathogenic mechanisms in centronuclear myopathies. Front Aging Neurosci. 2014;6:339.

55. Shieh PB, Kuntz NL, Dowling JJ, Muller-Felber W, Bonnemann CG, Seferian AM, et al. Safety and efficacy of gene replacement therapy for X-linked myotubular myopathy (ASPIRO): a multinational, open-label, dose-escalation trial. Lancet Neurol. 2023;22(12):1125–39.

