## Supplemental Figure 1 and Tables 1-2 for "A scalable and equitable framework for target and patient prioritisation in rare disease antisense therapeutics"

### Supplementary Figures

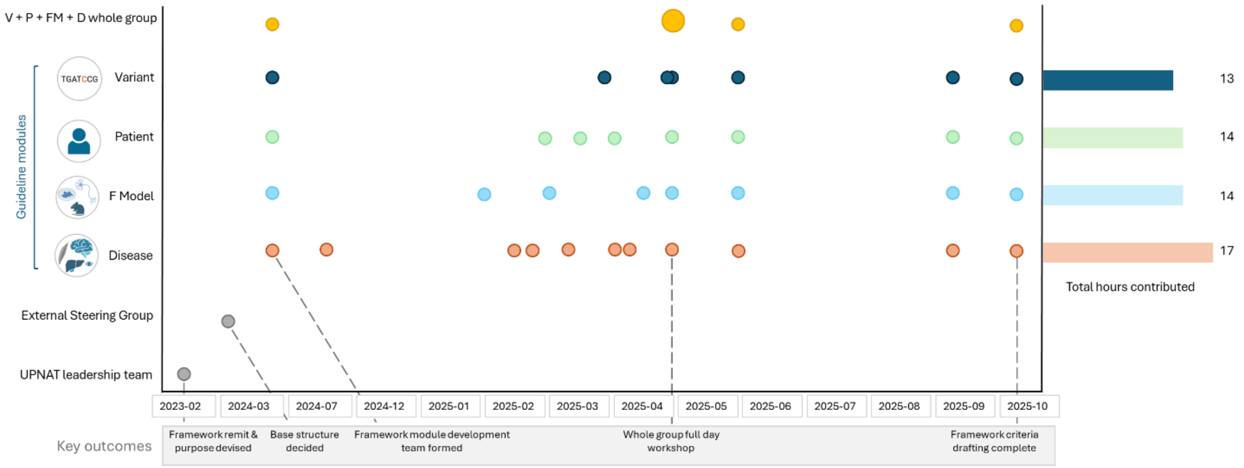


**Supplementary Figure 1**: **Timeline of framework development.** Key dates in the framework development timeline, from initiation (2023-02) to a complete draft (2025-10). Dates and key outputs are described along the X axis, the contributing group is indicated on the Y axis and cumulative time spent on development for each module is shown to the right. The UPNAT leadership team consisted of six individuals, the external steering group was a multidisciplinary group of >30 expert scientists and clinicians with diverse NAT and rare disease expertise from across the UK, and the development leadership group consisted of nine multi-disciplinary colleagues split across four modules. This group was formed to achieve two pre-defined aims: 1) generating guidance for the selection of patients and targets for RNA therapies in the UK, and 2) facilitating implementation of this framework in select highly specialised services as exemplar.

#### Supplementary Tables

**Supplementary Table 1: Expert group selected to develop the UPNAT target selection framework.**

| Individual | Designated section | Role | Institute/Company | Relevant expertise | Expert category |
|---|---|---|---|---|---|
| Dr Emma Clement | Disease | Leader | Great Ormond Street Hospital | Consultant in Clinical Genetics and Genomics Medicine specialising in paediatric rare disease with a focus on genetic deafness and deep knowledge of disease mechanisms and clinical presentations relevant to ASO therapy prioritisation. | Rare disease expert clinician, Clinical geneticist |

| | | | | | |
|---|---|---|---|---|---|
| Dr Hannah Titheradge | Disease | Leader | Birmingham Women's Hospital | Consultant clinical geneticist and Rare Disease lead for the Birmingham Women's Hospital with an in-depth clinical genetics background and experience in rare disease diagnoses, focusing on unmet medical need within NHS settings. | Rare disease expert clinician, Clinical geneticist |
| Prof Stephan Sanders | Functional model | Leader | Oxford University | Professor of Paediatric Neurogenetics and internationally recognised as an expert in human genetics, bioinformatics, and gene discovery, providing strong grounding in gene-level analysis. | Rare disease expert clinician, Academic molecular geneticist, Preclinical model development expert |
| Prof Jacqueline van der Spuy | Functional model | Leader | UCL | Professor of Molecular and Cellular Biology with specialist knowledge of inherited retinal dystrophies and experience in developing advanced functional models for fundamental science research and ASO preclinical development. | Academic molecular geneticist, Preclinical model development expert |
| Prof Carlo Rinaldi | Patient | Leader | Oxford University | Professor of molecular and translational neuroscience and Consultant Neurologist with extensive experience in translational research and clinical care of neurological diseases, providing expertise on patient-level feasibility and therapeutic applicability of ASOs. | Rare disease expert clinician, Academic molecular geneticist, Preclinical model development expert |

| | | | | | |
|---|---|---|---|---|---|
| Prof Manju Kurian | Patient | Leader | Great Ormond Street Hospital | Professor of Paediatric Neurogenetics and Consultant Paediatric Neurologist, internationally recognised for her work on childhood-onset neurogenetic disorders, bringing expertise in patient pathways, comorbidity assessment, and ethical considerations for early-phase ASO therapy. | Rare disease expert clinician, Academic molecular geneticist, Preclinical model development expert |
| Prof Jenny Taylor | Variant | Leader | Oxford University | Professor of Translational Genomics and Director the Oxford Biomedical Research Centre Genetics Theme, with expertise in variant interpretation, molecular diagnostics, and translational research, supporting rigorous variant-level assessment for ASO targeting. | Academic molecular geneticist, Preclinical model development expert |
| Dr Ana Lisa Tavares | Variant | Leader | Genomics England | Clinical Lead for Rare Disease research at Genomics England with specialist expertise in variant annotation and functional genomics, experienced in assessing variant pathogenicity and biological tractability in the context of precision therapeutic development. | Rare disease expert clinician, Clinical geneticist |
| Dr Nour Elkhateeb | Variant | Leader | Cambridge University Hospitals, Genomics England | Clinical Geneticist and Genomics England Research Fellow with expertise in genomics and transcriptomic, skilled in bioinformatic and laboratory-based evaluation of variant effects to inform their suitability for ASO intervention. | Rare disease expert clinician, Clinical geneticist |

| | | | | | |
|---|---|---|---|---|---|
| Dr Carme Camps | Variant | Developer | Oxford University | Molecular and computational biologist at Oxford University with expertise in genomic medicine and transcriptomics, combining bioinformatic and experimental approaches to interpret variant effects and assess their suitability for therapeutic intervention, including antisense oligonucleotide (ASO) strategies. | Academic molecular geneticist |

**Supplementary Table 2: Eleven market approved ASO therapies to calibrate and refine the UPNAT target selection framework.**

| **Drug name** | **Disease Target (MIM)** | **Gene Target** | **Assessor category** |
|---|---|---|---|
| Eteplirsen | Duchenne muscular dystrophy (310200) | *DMD* | Assessor 1: Academic molecular geneticist<br>Assessor 2: Preclinical model development expert<br>Assessor 3: Rare disease expert clinician |
| Nusinersen | Spinal muscular atrophy (clinical subtypes SMA 1-3) (253300, 253550, 253400) | *SMN2* | Assessor 1: Academic molecular geneticist<br>Assessor 2: Preclinical model development expert<br>Assessor 3: Rare disease expert clinician |
| Golodirsen | Duchenne muscular dystrophy (310200) | *DMD* | Assessor 1: Academic molecular geneticist<br>Assessor 2: Preclinical model development expert<br>Assessor 3: Rare disease expert clinician |
| Viltolarsen | Duchenne muscular dystrophy (310200) | *DMD* | Assessor 1: Academic molecular geneticist<br>Assessor 2: Preclinical model development expert<br>Assessor 3: Rare disease expert clinician |
| Casimersen | Duchenne muscular dystrophy (310200) | *DMD* | Assessor 1: Academic molecular geneticist<br>Assessor 2: Preclinical model development expert<br>Assessor 3: Rare disease expert clinician |

| | | | |
|---|---|---|---|
| Tofersen | Amyotrophic lateral sclerosis (SOD1 related) (105400) | *SOD1* | Assessor 1: Rare disease expert clinician, Academic molecular geneticist, preclinical model development expert<br>Assessor 2: Academic molecular geneticist |
| Eplontersen | Hereditary transthyretin amyloidosis polyneuropathy (105210) | *TTR* | Assessor 1: Academic molecular geneticist<br>Assessor 2: HCPC clinical scientist |
| Inotersen | Hereditary transthyretin amyloidosis polyneuropathy (105210) | *TTR* | Assessor 1: Academic molecular geneticist<br>Assessor 2: HCPC clinical scientist |
| Olezarsen | Familial chylomicronemia syndrome | *APOC3* | Assessor 1: Academic molecular geneticist<br>Assessor 2: HCPC clinical scientist |
| Volanesorsen | Familial chylomicronemia syndrome | *APOC3* | Assessor 1: Academic molecular geneticist<br>Assessor 2: HCPC clinical scientist |
| Donidalorsen | Hereditary angioedema | *KLKB1* | Assessor 1: Academic molecular geneticist<br>Assessor 2: HCPC clinical scientist |

**Supplementary Table 3: Application of the UPNAT framework across four illustrative patient case examples**

**Supplementary Tables 4: UPNAT disease assessment module.**

**Supplementary Tables 5: UPNAT functional model assessment module.**

**Supplementary Tables 6: UPNAT patient assessment module.**

**Supplementary Table 7: Variant module ASO amenability assessments for the patient cohort.**
