## Supplemental Tables 4-6 for "A scalable and equitable framework for target and patient prioritisation in rare disease antisense therapeutics"

Supplementary Table 4: UPNAT disease assessment module.

| Criteria category | Explanation | Criteria | Score to support | Weighting |
|---|---|---|---|---|
| **Step 1: Exclusion** | | | | |
| **Expected treatment outcome** | If no clinical benefit is predicted then this disease should not be considered for ASO development. This would include for example a congenital onset disease with no modifiable aspect to be targeted by ASO treatment or where the anticipated treatment window has passed, for example a growth plate fusion in a skeletal dysplasia. | No clinical benefit predicted. | Exclusion | NA |
| **Alternative treatment options** | A successful treatment option addressing the underlying cause of disease or successfully managing symptoms, negating the need for novel therapy development, is available for disease. This may be outside the UK and, in this case, resources need to be directed to assess this option for UK patients and make it available within the UK. Ideally, this treatment option is available within the UK and available through the NHS. However, if assessing for a named patient, individual patient treatment success must be considered and this may not exclude the disease (please see patient module). | Alternative successful treatment or symptom management is available. | Exclusion | NA |
| **Target tissue** | No evidence for ASO targeting to disease tissue at this time. Currently, we recommend those tissues listed under categories 1-3 below and any others for which identifiable tissue targeting data is available. | No evidence for ASO targeting to disease tissue at this time. | Exclusion | NA |
| **Step 2: Scoring** | | | | |
| **Target tissue** | Category 1: Strong evidence of tissue targeting and high efficacy upon delivery. | Category 1 tissues could include liver (Olezarsen), CNS (Nusinersen) and eye (Fomiversen). | 3 | 1 |
| | Category 2: Strong evidence of tissue targeting but lower efficacy upon delivery. | Category 2 tissue could include muscle (Eteplirsen). | 2 | 1 |
| | Category 3: Preliminary data available to support tissue targeting. | Category 3 tissue could include colon (Oblimersen). | 1 | 1 |
| **Mechanism of action** | To successfully develop an ASO, disease mechanism of action must be known either fully or mostly understood. | The underlying mechanism of disease is understood. | 3 | 1 |
| | A partially understood disease mechanism could still provide enough understanding for ASO development. | The underlying mechanism of disease is partially understood. | 2 | 1 |
| **Natural history** | Strong knowledge of disease course based upon longitudinal studies of gene specific cohorts ensures a comprehensive understanding of disease and offers the most promising disease groups for novel drug development. | Natural history study of disease with a gene specific cohort longitudinally tracked. | 3 | 1 |

| | | | | |
|---|---|---|---|---|
| | Moderate knowledge of expected disease progression and outcome based on disease/gene specific cohorts. This could include natural history studies of smaller disease/gene specific cohorts with some longitudinally tracked data or larger cohorts with non-longitudinal data providing moderate confidence in disease characteristics and progression. Consideration can be given to discovery reports with sufficient evidence supporting a robust disease-gene association even if not currently classified as OMIM morbid. | Moderate knowledge of expected disease progression and outcome based on disease/gene specific cohorts. | 2 | 1 |
| **Clinical endpoints** | If a clinical measure (e.g., seizures and walk tests) and biomarker (e.g., inflammatory markers and metabolites) or surrogate biomarker is available for disease, this is ideal for ASO development. | Clinical measure and biomarker are available for disease. | 3 | 1 |
| | To properly assess ASO impact, clinical measures are essential and could be assessed alongside patient reported clinical outcomes. Biomarkers (inclusive of surrogate biomarkers) are quantifiable characteristics of biological processes and can be used to measure disease. However, they do not always correlate with patient experience or clinical state. Having both is ideal but this is not always possible. | There is an established clinical measure or biomarker of disease severity available. | 2 | 1 |
| | There may be a biomarker available which is not established but can contribute to tracking disease severity. | There is a clinical measure or biomarker available that can be used but it is not well established. | 1 | 1 |
| **Clinical trials** | Previous clinical trials suggest important information, such as biomarkers and disease models, are available for disease. It should be noted that, if failed, assessors should be aware of the reason and proceed appropriately. | Regardless of success and drug modality, a clinical trial has been conducted for this disease. | 2 | 1 |
| **Tissue biopsy** | Access to disease tissue for biopsy for *ex vivo* ASO efficacy assessment is beneficial for ASO development. Inability to access disease tissue for biopsy cannot exclude a disease as many disease tissues are not directly accessible and models can be developed in the absence of biopsy. If the target gene is ubiquitously expressed, an alternative biopsy, such as skin fibroblast, could be acceptable. | The disease tissue is accessible by biopsy. | 3 | 1 |
| **Reduced weighting criteria** | | | | |
| **Alternative treatment options** | If no treatment options are available, then there is an urgent need for novel therapies for patients with this disease. | There is no successful treatment option available for this disease. | 3 | 0.5 |
| | There may be a treatment or management option for symptoms of disease with partial success (targeting one aspect of disease or slowing progression only) and/or significant side effects or morbidity (this may include side effects from treatment or method of delivery). | Treatment or symptom relief is possible but not adequate. | 2 | 0.5 |
| **Disease prevention** | The primary goal of ASO treatment is to halt the progression of disease however in some cases, disease prevention or reversal could be possible. For example, a highly penetrant disease with a presymptomatic period. In these disease cases, ASO intervention is highly attractive as a therapeutic option. | Disease resolution is possible from intervention. | 3 | 0.5 |

| | | | | |
|---|---|---|---|---|
| **Disease severity** | A life-threatening disease has high demand for successful treatment options for patients. Similarly, a severely debilitating disease (impact on patient day-to-day functioning) has high demand for successful treatment options for patients. | Disease is life threatening or severely debilitating. | 3 | 0.5 |
| **Partial or complete disease targeting** | Therapy expected to target the entirety of disease. Disease characteristics can favour an ASO intervention strategy that enables full, or near complete, targeting of disease. For example, a single organ is involved in disease or, if multisystemic, one organ is the root cause of other organ system engagement and targeting this one organ addresses all disease characteristics. Cases such as these are favourable to ASO treatment. | Full disease, or near complete, targeting possible with ASO intervention. | 3 | 0.5 |
| | In some disease cases, it will not be possible for ASO treatment to target the full disease spectrum and therefore the intervention needs to be well thought out with the patient in mind. An example of this would be a multisystemic disease without a single causative organ linked to the full disease spectrum. | Partial disease targeting possible with ASO intervention. | 2 | 0.5 |

Supplementary Table 5: UPNAT functional model assessment module.

| Criteria category | Explanation | Criteria | Score to support | Weighting |
|---|---|---|---|---|
| **Step 1: Exclusion** | | | | |
| **Gene expression and protein levels** | The assessor must understand which tissues and cell types express the gene in order to select an appropriate experimental model. Ideally, both the spatial and temporal patterns of gene expression should be considered. | Spatial and/or temporal gene expression is unknown, thereby preventing suitable model selection for ASO development. | Exclusion | NA |
| **Step 2: Scoring** | | | | |
| **Gene expression and protein levels** | Gene is known to be expressed (RNA) or detectable (protein) in the specific cell type being targeted for ASO treatment. | Gene is expressed in the cell type being targeted for ASO treatment. | 3 | 1 |
| **Gene expression and protein levels - screening experimental models** | The gene is expressed in a simple in vitro human model (e.g. HEK293, K562, SH-SY5Y) likely to be chosen for rapid functional investigation for ASO development and toxicity assessment. | Gene is expressed in a simple in vitro human model. | 3 | 1 |
| **Gene expression and protein levels - lead optimisation experimental models** | The gene is expressed in a functionally relevant in vitro human model (e.g., NGN2 neurons) for functional investigation for ASO development. | Gene is expressed in a functionally relevant in vitro human model. | 3 | 1 |
| **Gene expression and protein levels - human preclinical experimental models** | The gene is endogenously expressed in a functionally relevant human in vitro tissue or organoid model (e.g., contracting muscle, retinal organoid). | Gene is endogenously expressed in a functionally relevant human in vitro tissue or organoid model. | 3 | 1 |
| **Gene expression and protein levels - nonhuman preclinical experimental models** | The gene is expressed in a functionally relevant non-human in vivo model (e.g., UBE3A-AS) and the target sequence is shared with humans (e.g., conserved element or humanised model). | Gene is expressed in a functionally relevant non-human in vivo model with shared target sequence with humans. | 3 | 1 |
| **Gene expression and protein levels - nonhuman preclinical experimental models** | The gene is expressed in a functionally relevant non-human in vivo model (e.g., UBE3A) but sequence differs from human. | The gene is expressed in a functionally relevant nonhuman in vivo model (e.g., UBE3A) but sequence differs from human. | 1 | 1 |
| **Gene function** | Gene has a well-established function within the cell type being studied for ASO development. | Gene function within relevant cell type known. | 3 | 1 |
| **Gene function - functional assays** | Well established assays to study the function of a gene enable assessment of ASO efficacy. | Well established assays for study of gene function are available. | 3 | 1 |
| **Previous drug development** | If an ASO intervention has been developed for a gene already and shown to work, it supports that the gene is | ASO intervention has successfully been developed for this gene previously. | 3 | 1 |

| | | | | |
|---|---|---|---|---|
| | amenable to ASO treatment and further strategies can be developed. | | | |
| **Previous drug development** | Gene has been studied in a therapeutic context. | Gene has been studied in a therapeutic context. | 2 | 1 |

Supplementary Table 6: UPNAT patient assessment module.

| Criteria category | Explanation | Criteria | Score to support | Weighting |
|---|---|---|---|---|
| **Step 1: Exclusion** | | | | |
| **Life expectancy** | If patient is at such an advanced or critical disease stage that the potential benefits of ASO therapy are outweighed by the risk, ASO treatment is not appropriate for this patient. | Patient is in an advanced or critical stage of disease and the risk of ASO therapy outweighs perceived benefit. | Exclusion | NA |
| **Patient prognosis** | Provided that an ASO is not readily available and taking into account the time taken to develop an ASO therapy (minimum 2 years), does the benefit outweigh risk of ASO treatment for a patient within this time frame? If risk outweighs benefit for the patient, they should be excluded. | With time taken to develop the ASO, the benefits for patient no longer outweigh the risk. | Exclusion | NA |
| **Comorbidity** | If a patient has a comorbidity requiring a treatment that cannot be stopped, which prevents the development of ASO therapy, then the patient will not be a candidate. | Patient has a comorbidity with treatment which may impact ASO therapy. It is not possible to remove patient from comorbidity treatment. | Exclusion | NA |
| | If a patient has a comorbidity with a severe clinical course and prognosis then ASO development for the less severe disease is not the priority for the patient. | Patient has a comorbidity with a clinical course predicted to be severe and life threatening. | Exclusion | NA |
| | Patient has a state or condition (e.g. pregnancy or severe coagulopathy), which excludes them from ASO treatment at time of assessment. | Patient has an excluding state or condition. | Exclusion | NA |
| **Complications within disease** | Complications of disease are present within the patient, which excludes them from ASO treatment (e.g. severe scoliosis for intrathecally administered ASOs). | Patients have complications related to their disease that could exclude them from ASO treatment. | Exclusion | NA |
| **Toxicity** | Standard measurements (e.g. full blood count, LFTs, EEG, ECG) may need to be taken for toxicity assessment. If the patient cannot tolerate this or becomes contraindicated over the course of the treatment, toxicity can't be properly assessed. | Patient cannot have standard measurements taken for toxicity assessment and therefore must be excluded. | Exclusion | NA |
| **ASO tolerability** | Thrombocytopenia (low platelet count) has been reported as an adverse effect of ASO therapies. Platelet count and other blood safety markers should be monitored during treatment. Given current knowledge of ASO adverse effects, all markers should be monitored. If the patient has an increased risk of thrombocytopenia, ASO treatment should be carefully considered. | Patient has an increased risk of thrombocytopenia. | Exclusion | NA |
| | Communicating hydrocephalus has been reported as an adverse effect of ASO administration via intrathecal, intracerebral or intraventricular injection in patients. The risk of an individual patient to this adverse effect should be considered if ASO is to be delivered via these routes. If treated, the patient could be included. | If the ASO is to be delivered via intrathecal, intracerebral or intraventricular injection to a patient who has hydrocephalus at baseline, they should be excluded from ASO therapy. | Exclusion | NA |

| | | | | |
|---|---|---|---|---|
| | Kidney toxicity has been indicated across several ASO drugs delivered systemically (such as nusinersen, inotersen, golodirsen, viltolarsen and casimersen). Pre-existing kidney dysfunction in a patient requiring systemic ASO delivery needs careful assessment as to whether the benefits outweigh the risks. | A patient requiring systemic ASO delivery with abnormal kidney function or predisposition to kidney complications must be more carefully considered for ASO treatment. If risk outweighs benefit, patient should be excluded. | Exclusion | NA |
| | Hepatotoxicity has been indicated across mipomersen and inotersen ASO therapies. Pre-existing liver dysfunction in a patient requiring ASO delivery to the liver needs careful assessment as to whether the benefits outweigh the risks. | A patient requiring delivery of an ASO to the liver with abnormal liver function or predisposition to liver complications must be more carefully considered for ASO treatment. If risk outweighs benefit, patient should be excluded. | Exclusion | NA |
| | Clinical deterioration of patients receiving CSF administered (intrathecal, intracerebral or intraventricular) ASO has been marked with a change in routine CSF parameters including increase in opening pressure. Patients should have regular monitoring of CSF parameters for safety. If a patient presents pre-treatment with abnormal inflammatory markers in the CSF and/or increase in opening pressure, risk of ASO treatment needs careful discussion. | A patient requiring CSF delivery of an ASO with abnormal CSF routine parameters including increased CSF opening pressure pre-treatment must be more carefully considered for ASO treatment. If risk outweighs benefit, patient should be excluded. | Exclusion | NA |
| **ASO administration** | The patient may have a device or disorder which prevents specific administration routes (e.g. presence of an implantable CNS device which interferes with repetitive intrathecal delivery). If this is the only route available to deliver the ASO, then the patient cannot be considered for ASO therapy. | The specific ASO administration route is not achievable for the patient. | Exclusion | NA |
| **Target engagement** | For some diseases, a patient biopsy (or relevant disease measurement) may be needed for target engagement before, during or after treatment. The ability to take a biopsy from a tissue would be beneficial to assess ASO treatment. If the patient would not tolerate repetitive biopsy they cannot be prioritised for ASO development. | Patient would either tolerate a single biopsy (or relevant disease measurement) only or cannot tolerate biopsy. | Exclusion | NA |
| **Monitoring efficacy** | Patients for which the clinically meaningful endpoints or measurements of biomarkers of efficacy are accessible present a more favourable target for ASO development and treatment. | Clinically meaningful endpoint or measurement of biomarkers of efficacy accessible for this patient | 3 | 1 |
| **Patient prognosis** | If an ASO is readily available for a given disease and, at this time point, the benefit outweighs the risk of ASO treatment for the patient, use of this developed ASO should be pursued. | An ASO is currently available and the benefit of use for the patient outweighs the risk. | 3 | 1 |
| | An ASO may be in development and suitable for the patient in question. If the benefits outweigh the risks, given time remaining in development, treatment should be pursued. | An ASO is in development and the benefit of use for the patient outweighs the risk. | 2 | 1 |
| | Taking into account the time taken to develop an ASO therapy, does the benefit outweigh risk of ASO treatment for a patient? If the benefit outweighs the risk, this patient should be considered for ASO development. | With time taken to develop the ASO, the benefits for patient still outweigh risk. | 1 | 1 |
| **Comorbidity** | A patient with no comorbidities (or secondary features of the disease) is an ideal candidate for novel therapy development. | Patient has no comorbidities (or secondary features of the disease). | 3 | 1 |

| | | | | |
|---|---|---|---|---|
| | Patients may present with comorbidities (or secondary features of the disease) that are not predicted to impact clinical progression of patient and so do not exclude patient. | Patient has a comorbidity (or secondary features of the disease) which is not predicted to impact on their clinical progression. | 2 | 1 |
| | A patient may have to be removed from an unrelated treatment for ASO therapy development. If this is the case, the benefit to risk of treatment alterations must be carefully considered. | Patient has a comorbidity (or secondary features of the disease) which is treated and which could impact on ASO therapy. | 1 | 1 |
| **Conventional treatment response** | A treatment option is available for the disease however, if the patient is not responding to the treatment, and the benefit of an ASO treatment outweighs the risk, the patient should be prioritised for an ASO treatment. | Patient is not responding to conventional treatment option available for disease. | 1 | 1 |
